# Descriptives and genetic correlates of eating disorder diagnostic transitions and presumed remission in the Danish registry

**DOI:** 10.1101/2024.09.05.24313142

**Authors:** Mohamed Abdulkadir, Janne Tidselbak Larsen, Loa Clausen, Christopher Hübel, Clara Albiñana, Laura M. Thornton, Bjarni J. Vilhjálmsson, Cynthia M. Bulik, Zeynep Yilmaz, Liselotte Vogdrup Petersen

## Abstract

**Objective:** Eating disorders (EDs) are serious psychiatric disorders with an estimated 3.3 million healthy life-years lost worldwide yearly. Understanding the course of illness, diagnostic transitions and remission, and their associated genetic correlates could inform both ED etiology and treatment. The authors investigated occurrences of ED transitions and presumed remission and their genetic correlates as captured by polygenic scores (PGSs) in a large Danish register-based cohort.

**Methods:** The sample compromised of 10,565 individuals with a diagnosis of anorexia nervosa (AN), bulimia nervosa (BN), or eating disorder not otherwise specified (EDNOS) with at least two registered hospital contacts between 1995 and 2018. Based on medical records, occurrence of diagnostic transitions and periods of presumed remission were identified. Associations between 422 PGS and diagnostic transitions and presumed remission were evaluated using Cox proportional hazard models.

**Results:** A minority of ED cases (14.1%-23.1%) experienced a diagnostic transition. Presumed remission ranged between 86.9%-89.8%. Higher (one SD increase) PGS for major depressive disorder and multisite chronic pain were positively associated with transitioning from AN to either BN or EDNOS. Higher PGS on a measure of body fat percentage and financial difficulties were positively associated with presumed remission from AN. Higher PGS for mood swings was positively associated with presumed remission from EDNOS whereas higher PGS for health rating showed the opposite.

**Conclusions:** The authors found that most ED patients did not experience diagnostic transitions but were more likely to experience a period of presumed remission. Both diagnostic transitions and presumed remission have significant polygenic component.

## Introduction

Eating disorders (EDs) are serious psychiatric disorders which can have a complicated course of illness characterized by diagnostic transition from one ED to another, and eventually remission for some patients (1–6). Annually, it is estimated that 3.3 million healthy life years are lost worldwide due to anorexia nervosa (AN), bulimia nervosa (BN), and eating disorder not otherwise specified (EDNOS) (7). Studies have reported varying frequencies of diagnostic transitions across EDs. For instance, some case-cohort and clinical studies found that the transition from AN to BN occurs in 13%-54% of cases, and from BN to AN in 7%-27% of cases (1–3,6). However, a large Swedish population register study (8) reported much lower transition probabilities (2%) from AN to BN and vice versa, with the transition to EDNOS occurring in 17%-24% of AN cases and in 18%-22% of BN cases (3,8). Estimates of ED remission also differ in the literature, ranging from 27%-78% for AN cases, 40%-83% for BN cases, and about 49% of EDNOS cases in studies with a minimum follow-up of 6 years (2,5,8). These differences could be due to variations in study design, assessment methods, or follow-up time.

It is unclear what factors determine a patient’s risk of transitioning from one ED to another or their duration of illness. Previous research suggests that greater experience of psychopathology (e.g., anxiety) may increase the likelihood of transition from AN to BN (1,6,9). Anthropometric factors such as body mass index (BMI) may also play a role as individuals who transition from AN to BN tend to have higher current, past minimum, and past maximum BMIs compared to those who do not transition (9). Similarly, higher BMI and traits such as premorbid perfectionism, state anxiety, and trait anxiety are associated with lower occurrence of ED remission (10–12).

One interesting approach to understanding biological factors that may influence ED transition and remission is to explore genetic factors that contribute to the risk of EDs (4,13). Most genome-wide association studies (GWAS) of EDs have focused on AN (14–17) and have not examined genetic factors influencing diagnostic transition or remission. Polygenic scores (PGSs) offer a powerful method to leverage the small-to-moderate effect sizes of common single nucleotide polymorphisms (SNPs) identified in a GWAS into a single continuous score that surpasses the explanatory power of individual SNPs (4). This approach has been successful in exploring the genetic relationship between traits by associating the PGS for one trait (e.g., BMI) with a phenotypic measurement of another trait (18).

Therefore, the aims of our study were two-fold: (i) quantify occurrences of ED transitions and remission; and (ii) investigate whether diagnostic transitions and ED remission are associated with PGSs of various complex traits in a nationwide sample of treatment-seeking individuals in Denmark (19).

## Methods and Materials

### Study population

Using data from the Danish National Patient Register (NPR) (20) and the Psychiatric Central Research Register (PCRR) (21), the study population comprised individuals: (i) born in Denmark between May 1, 1981 and December 31, 2009; (ii) who were alive and living in Denmark at their 6^th^ birthday; (iii) who received an International Classification of Diseases, 10^th^ edition (ICD-10) primary or secondary diagnosis for AN (F50.0, F50.1), BN (F50.2, F50.3), or EDNOS (F50.8, F50.9) on or after their 6^th^ birthday; (iv) for whom genotype data were available; (v) who had at least two registered ED-related hospital contacts between 1995 and 2018 after the age of 6 years. Genotype data for EDNOS cases without any other lifetime ED diagnosis were obtained from the iPSYCH2015 case-cohort sample (also including the iPSYCH2012 case cohort) (19,22), for AN cases from the Danish branch of the Anorexia Nervosa Genetics Initiative (ANGI-DK) (23), and for BN cases from the Danish branch of the Eating Disorders Genetics Initiative (EDGI-DK) (24). In total, we included 10,565 individuals with EDs, with their first recorded diagnosis being: AN, n = 6,901; BN, n = 1,417; EDNOS, n = 2,247. Each individual was followed up until December 31, 2018. Sex information was obtained from the NPR and PCRR. Sex is assigned at birth; however, each individual over the age of 18 years can legally change their sex information in the registers based on their gender identity. Therefore, from here onwards we report on gender. Ancestry was based on genetic information and was determined by calculating robust Mahalanobis (squared) distances using the first two genomic principal components. This approach relies on the fact that the Danish population is fairly homogenous (22) and calculating robust distance captures individuals with an European ancestry.

### Outcome data

#### ED episodes

From the inpatient and outpatient hospital contacts in the NPR and the PCRR, we defined distinct ED episodes for each individual (for detailed information, see **Supplemental Methods** and **Figure S1**). First, we combined patient records into distinct hospital contacts. We further merged contacts into ED episodes (i.e., a time period in which an individual had a diagnosis of an ED). The merging of records into contacts and episodes resulted in three possible ED episodes: AN; BN; or EDNOS. We also included an episode of presumed remission; individuals that were contact free regarding their ED for at least 2 years.

#### Time to diagnostic transition

Based on the defined ED episodes, we first grouped individuals by their first ED diagnosis. Second, we defined a diagnostic transition as an individual receiving another ED diagnosis in a subsequent ED episode. Time to diagnostic transition was calculated as the difference between the beginning of the first ED episode and the beginning of the episode in which the individual received a different ED diagnosis.

#### Time to presumed remission

Time to presumed remission was calculated as the time difference between the moment the person is at risk for presumed remission (two years following the start of the first episode) and the date the individual met the above-mentioned criteria for presumed remission.

### Genetic data

#### Genotyping

Dried blood spot samples were obtained through routine screening for congenital disorders from nearly all Danish newborns after 1981 and stored at the Danish Newborn Screening Biobank (25). Genotyping of the DNA extracted from the blood spot samples was carried out and described elsewhere (19,22,23,25). This study was approved by the Danish Data Protection Agency, the Danish Scientific Ethics Committee, the Danish Health Data Authority, and the Danish Newborn Screening Biobank Steering. The Danish Scientific Ethics Committee, in accordance with Danish legislation, has, for this study, waived the need for informed consent in biomedical research based on existing biobanks (19).

#### PGSs calculations

PGSs were calculated for ED cases with genetic data (n cases = 10,565) using LDpred2 (26) for 940 traits and with the meta-PRS (27) approach for 6 traits (i.e., attention deficit hyperactivity disorder [ADHD], AN, autism spectrum disorder, bipolar disorder, major depression, and schizophrenia). We performed pre-filtering on 940 LDpred2-calculated PGSs (e.g., duplicates and highly correlated PGSs; for details see **Supplemental Methods** and **Figure S2**). After filtering, a total of 422 PGSs remained for analyses and were standardized (**Table S1**).

### Statistical analyses

We tested the association between PGSs (one at a time) and our outcome measures using Cox proportional hazard models. We did not exclude individuals based on genetic ancestry and instead controlled for population stratification using the first five genomic principal components as covariates in the Cox regression models. We did not include transitions from BN to AN or EDNOS or from EDNOS to AN or BN in our PGS analysis due to the infrequent occurrence of these transitions in our sample (**Table 1**). We estimated hazard ratios with 95% confidence intervals for each outcome for one SD increase in the PGS. We corrected for the number of PGSs tested (n = 422) by calculating the number of effective tests using the *meff()* function (PoolR package) on the genetic correlation matrix of the selected 422 PGS applying the Galwey method (28). The number of effective tests was estimated to be 317, therefore the threshold for multiple testing was set at α < 1.57 × 10^-4^.

**Table 1:** Descriptive statistics of the sample by first eating disorder diagnosis presentation.

|  | **Total sample**<br>**(n = 10,565)** |  | **AN as first episode**<br>**(n = 6,901)** |  | **BN as first episode**<br>**(n = 1,417)** |  | **EDNOS as first episode**<br>**(n = 2,247)** |  |
| --- | --- | --- | --- | --- | --- | --- | --- | --- |
| **Binary descriptives** | **n** | **%** | **n** | **%** | **n** | **%** | **n** | **%** |
| Gender, women | 9,784 | 92.6 | 6,425 | 93.1 | 1,381 | 97.5 | 1,978 | 88.0 |
| European ancestry | 9,037 | 85.5 | 5,952 | 86.2 | 1,201 | 84.8 | 1,884 | 83.8 |
| Transition to BN |  |  | 370 | 5.4 |  |  | 153 | 6.8 |
| Transition to EDNOS |  |  | 661 | 9.6 | 175 | 12.3 |  |  |
| Transition to AN |  |  |  |  | 165 | 11.6 | 404 | 17.8 |
| Transition to BN or EDNOS |  |  | 968 | 14.1 |  |  |  |  |
| Transition to AN or EDNOS |  |  |  |  | 309 | 21.8 |  |  |
| Transition to AN or BN |  |  |  |  |  |  | 521 | 23.1 |
| Presumed remission | 9,273 | 87.7 | 5,999 | 86.9 | 1,273 | 89.8 | 2,001 | 89.0 |
| Relapsed from presumed remission | 1,585 | 15.0 | 970 | 14.0 | 290 | 20.5 | 325 | 14.5 |
| **Continuous descriptives** | **mean** | **sd** | **mean** | **sd** | **mean** | **sd** | **mean** | **sd** |
| Age at first diagnosis in years | 18.1 | 4.5 | 17.3 | 4.1 | 20.9 | 4.3 | 18.7 | 5.1 |
| Follow-up time in years | 8.9 | 5.2 | 9.0 | 5.4 | 9.1 | 4.9 | 8.2 | 4.9 |
| Time to transition to BN in years |  |  | 4.5 | 3.3 |  |  | 3.6 | 3.1 |
| Time to transition to EDNOS in years |  |  | 4.7 | 3.8 | 4.6 | 3.6 |  |  |
| Time to transition to AN in years |  |  |  |  | 4.0 | 3.4 | 2.6 | 2.5 |
| Time to transition to BN or EDNOS in years |  |  | 4.5 | 3.5 |  |  |  |  |
| Time to transition to AN or EDNOS in years |  |  |  |  | 4.0 | 3.2 |  |  |
| Time to transition to AN or BN in years |  |  |  |  |  |  | 2.7 | 2.7 |
| Time to presumed remission in years | 1.6 | 1.8 | 1.7 | 1.9 | 1.7 | 1.76 | 1.2 | 1.6 |
| Duration presumed remission in years | 5.3 | 4.7 | 5.4 | 4.9 | 4.9 | 4.3 | 5.0 | 4.3 |

AN, anorexia nervosa; BN, bulimia nervosa; EDNOS, eating disorder not otherwise specified; SD, standard deviation.

## Results

### Sample description

#### Demographics

We included a total of 10,565 (92.6% women; 85.5% European ancestry) individuals with genotype and ED diagnostic data in our analyses (**Table 1**): 6,901 individuals with AN, 1,417 individuals with BN, and 2,247 individuals with EDNOS as their first episode. The mean age at first diagnosis for AN, BN, and EDNOS in our sample were 17.3 (standard deviation [SD] = 4.1), 20.9 (SD = 4.3), and 18.7 (SD = 5.1) years, respectively. The mean follow-up time was 8.9 years (SD = 5.2).

#### Transitions

Among individuals with AN as their first episode, 14.1% transitioned to another ED with such transition taking an average time of 4.5 years (**Table 1**). The transition from AN to BN occurred in 5.4% of individuals, with an average time to transition of 4.5 years. A higher percentage of individuals with AN (9.7%) transitioned to EDNOS, with a mean time to transition of 4.7 years. Among individuals with BN as their first episode, 21.8% individuals transitioned to AN or EDNOS: specifically, 11.6% transitioned to AN and 12.3% transitioned to EDNOS, with an average transition time of 4 years and 4.6 years, respectively. Individuals presenting with EDNOS as their first episode showed the highest transitions; 23.1% of the individuals transitioned to AN or BN with a mean transition time of 2.7 years. More EDNOS cases transitioned to AN (17.8%) than to BN (6.8%).

#### Presumed remission

Regardless of ED diagnosis at first episode, 87.7% of individuals experienced a period of presumed remission with a mean time to presumed remission of 1.6 years (SD = 1.8; **Table 1**) with an average duration of the presumed remission period of 5.3 years (SD = 4.7). Presumed remission occurred in 86.9% of individuals with AN as first episode, with a mean time to presumed remission of 1.7 years (SD = 1.9). Among individuals with BN as first episode, 89.8% experienced a period of presumed remission with a mean time to presumed remission of 1.7 years (SD = 1.7). Of the individuals with EDNOS as their first episode, 89.0% experienced a period of presumed remission with a mean time to remission of 1.2 years (SD = 1.6). A minority (15%) of the cases experienced an ED diagnosis following a period of presumed remission; relapse was highest (20.8%) among those with BN diagnosis as their first episode.

### Association of PGSs with transition from AN to BN or EDNOS

Higher major depressive disorder (MDD) and higher multisite chronic pain PGS were significantly associated with a 15% greater hazard of transitioning from AN to BN or EDNOS, respectively (**Figure 1A**; see **Table S2** for full results).

**Figure 1:**
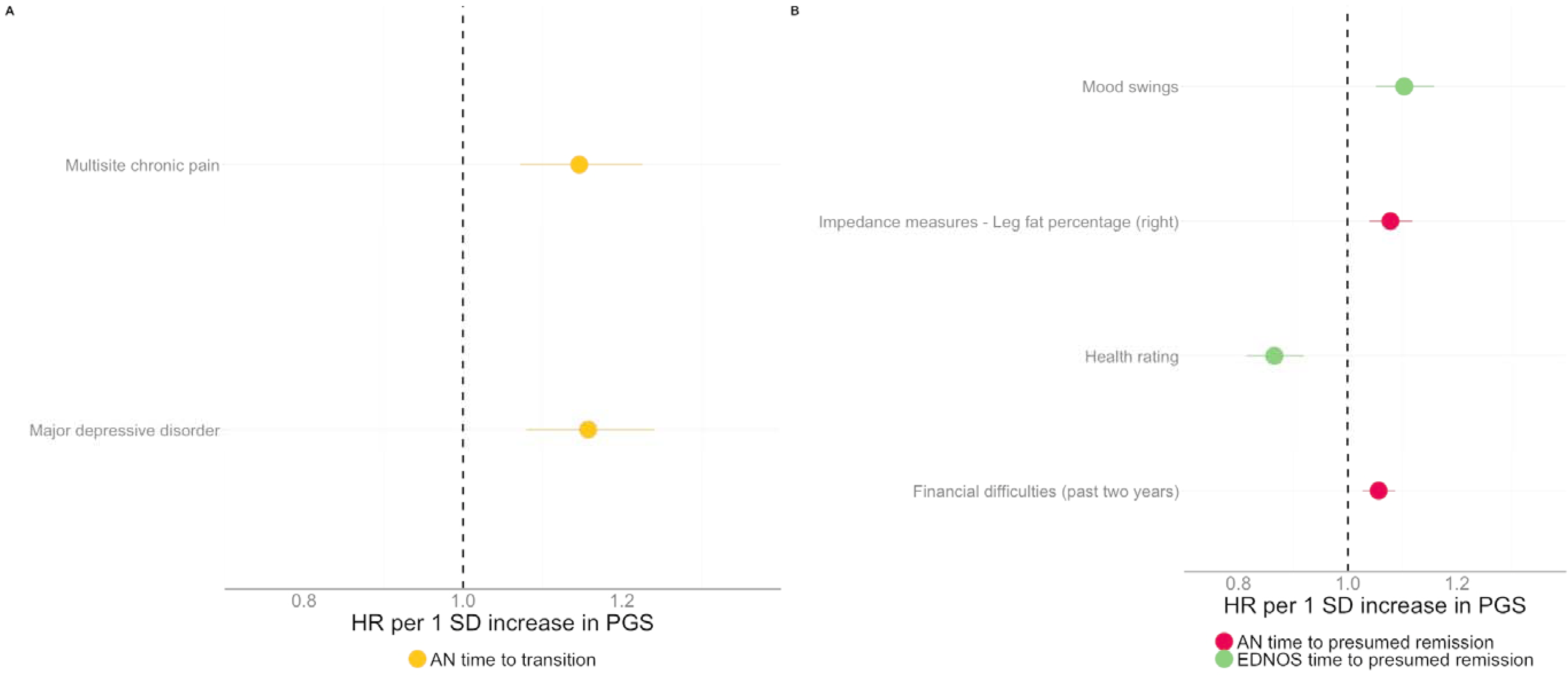
Associations between polygenic scores (PGS) and eating disorder (ED) diagnostic transition and remission. Depicted are the associations that remained significant after correcting for multiple hypothesis testing. Associations are depicted in hazard ratios (HR) per one standard deviation (SD) increase in the PGS with dots representing the HR point estimate and the line depicting the 95% confidence interval A. Associations of PGS with transitioning from AN to BN or EDNOS. B. Associations of PGS with ED presumed remission.

### Association of PGSs with time to presumed remission

Higher (one SD) PGS of a body fat percentage measure (i.e., leg fat percentage) was associated with an 8% greater hazard for presumed remission from AN (**Figure 1B; Table S3-S5**). In AN individuals, higher PGS on financial difficulties was also associated with a 5% greater hazard for presumed remission. Regarding EDNOS, each one SD increase in the PGS of overall health rating corresponded with a 12% lower hazard for presumed remission while a higher PGS on mood swings was associated with a 10% increased hazard for presumed remission.

## Discussion

In this Danish nationwide population study, we examined 10,565 individuals with EDs over an average follow-up time of 9 years. Overall, the occurrence of transitions (ranging between 5.4%-23.1%) in our sample fall on the lower end of previous estimates (1–3,6). For example, in a study in which AN cases were followed weekly, about a third of all AN cases transitioned to BN in a period of 7 years (2); their transition estimate is much higher compared to ours (5.4%). However, our estimate of transitions are more comparable to another registry-based study that found similar estimates that ranged between 2%-24% (8). There could be several explanations for our lower estimates of transitions. It could be that diagnostic transitions might occur in-between hospital visits and are therefore likely missed in register-based research. It is also possible that patients seek treatment elsewhere (e.g., private practice that is not recorded in the registries we utilized) or decide to no longer pursue further treatment. Despite differences in estimate of threshold ED transitions across studies, the general pattern to emerge is that diagnostic stability is the norm and if transitions do occur they often happen within the first five years of Illness (1–3,6,8).

Presumed remission was very likely in our sample; depending on the diagnosis of the first episode, between 86.9%-89.0% of the individuals experienced a period of presumed remission with only a minority (14.0%-20.5%) of the individuals experiencing relapse suggesting that this is a relatively stable state. The frequency of presumed remission in our study was comparable to partial recovery frequencies (78.4%-82.8%) reported in a longitudinal study of ED (2), suggesting that our definition likely reflects individuals that are on their way to recovery. However, our estimates for presumed remission are much higher compared to the only other register-based study on this topic that reported remission to vary between 23%-36% (8). The definition of remission in the other register-based study was defined as instances in which an ED diagnosis was no longer coded in the register at an assessment visit whereas ours was based on being contact free regarding ED for at least 2 years. The discrepancy in result is likely due to these differing operationalizations of a remission phenotype and might suggest that our estimates for presumed remission is an overestimation. However, the overall conclusion regarding remission in our study and those of the other register-based study (8) remain the same; remission is a more likely outcome than diagnostic transitions and remission is a fairly stable state.

Regarding our PGS analysis of transitions from AN to BN or EDNOS; higher PGS for MDD and multisite chronic pain (a phenotype defined as the sum of the number of body sites at which an individual reported chronic pain that lasted at least 3 months) corresponded with a 15% greater hazard of transitioning. Associations between AN and depression are well established with some of the shared etiology likely due to shared genomics; MDD is genetically positively correlated (*r*_g_ = 0.28, SE = 0.07, p = 8.95 × 10^−5^) with AN caseness (16,29,30). Our finding adds to the literature of AN and MDD and suggest that alleles that underlie MDD can also influence the course of AN. Along similar lines, multisite chronic pain is reported to be genetically negatively correlated (*r*_g_ = -0.06, SE = 0.03, p = 0.047) with AN (31). ED disturbances are elevated in patients with chronic pain, and findings from a longitudinal study show that adolescents with an ED are more likely to experience frequent or persistent pain (32). Furthermore, studies suggest that pain sensitivity may be decreased in individuals with AN and BN compared to non-ED controls (33–35). This change in pain perception is likely not due to deficits in interoception (sense of the physiological condition of the body (36)), which are implicated in both AN and BN (37). A recent study found no evidence for a mediation effect for interoception in the association between pain experience and EDs, which suggests that pain perception in AN may be altered through a different physiological system (37). One possible shared biological process that might explain the co-occurrence of chronic pain and ED could be central sensitization: a state in which the central nervous system responsiveness is increased to internal and external conditions, which is a process that is dysregulated in both conditions (32).

Regarding our PGS analysis and remission, higher PGS on a body fat measure (leg fat percentage) was associated with presumed remission from AN. This finding is of considerable relevance etiologically as the maintenance of low BMI and persistence of dietary restriction are unique and biologically challenging symptom profiles. How some individuals maintain a low BMI throughout their course of illness versus conceding biologically to increased caloric intake and weight gain may indeed rest in genetic susceptibility to both anthropometric and metabolic traits. Our findings suggest that those that harbor genetic variants that increases their bodyweight might counter the symptom profile (i.e., maintaining a low body weight) of AN and hence they contribute to presumed remission. We also report that higher PGS on financial difficulties was associated with remission from AN. Several longitudinal studies have implicated higher parental SES and increased risk of developing AN (38,40). Furthermore, AN is reported to be positively genetically correlated (r_g_ = 0.20 - 0.27) with several measures of educational attainment and a recent PGS study found that higher AN PGS is associated with higher SES in parents of AN cases (16,39). Our findings are likely captures the SES component of financial difficulties and might suggest that SES additionally plays a role in the course of AN.

Counter to our expectations, we found that higher PGS on overall health rating was negatively associated with remission from EDNOS. It is important to note that about 18% of the cases with EDNOS transitioned to AN suggesting that these could have been individuals that had not yet fulfilled criteria for AN at the time of their first assessment. It is therefore possible that the PGS on overall health rating captures the characteristics of this subgroup such as lower body weight and engagement in excessive exercise; both of which have been previously associated with a measure of overall health rating (41). If shown to true, this could potentially open a path in which the EDNOS group can be dissected into a more homogenous group using PGS. Unfortunately, our sample size was not sufficient to pursue this goal. Lastly, we also report a positive association between higher PGS on mood swings and remission from EDNOS. Mood swings are highly prevalent in EDNOS patients (43), and it is therefore likely that the clinical treatment for mood swings takes precedence which might appear as presumed remission from EDNOS.

The lack of significant associations between most of the tested PGSs and our outcomes could be due to several reasons. Despite the relatively large sample size, it is possible that our study lacked power to detect associations between the PGSs and transitioning from AN to BN or EDNOS, given the infrequent occurrences of these transitions (N cases that transitioned from AN to BN or EDNOS = 976). Furthermore, it is possible that both AN transitions and ED remission are largely determined by environmental factors (e.g., treatment). Lastly, it is also possible that our transition and remission phenotypes have a unique genetic architecture that is not best captured by our selection of PGS.

The strengths of this study include its large sample size and the long follow-up period compared to previous work (1–6). However, findings from this study should be interpreted in the context of certain limitations. In Denmark, hospital-based treatment is free of charge, lowering the barrier for individuals to seek treatment (19). However, we cannot rule out that sociodemographic factors could still have influenced treatment-seeking behavior. The diagnoses in the registers could have also been impacted by differing registration practices among hospitals departments and over time which could have impacted our estimates of diagnostic transition and remission (42). For example, a clinician might assign a diagnosis of EDNOS as a first diagnosis because threshold AN or BN is still under development. Similarly, a person with AN might on their way to recovery go through an EDNOS stage. In the previous examples, the transition to or from EDNOS is likely an artifact of diagnostic practice. Diagnostic practices when both AN and BN criteria are met can vary between hospitals as guidelines are not consistent across diagnostic manuals. However, in our work we have grouped hospital contacts into distinct ED episodes and episodes that are temporally close were merged into one episode which likely dampened the effects of such diagnostic practices. We can’t rule out that other diagnostic practices could have influenced our results. Our definition of presumed remission relied on being contact free for at least 24 months and is therefore not a direct measure of remission; hence why we refer to this phenotype as presumed remission. However, it is important to note that when individuals entered this period of presumed remission it lasted on average 5.3 years and only a minority (15%) of cases relapsed suggesting that our definition of presumed remission does approximate remission. Furthermore, it is important to reiterate that our study might have missed some diagnostic transitions as they could have occurred between hospital contacts. Genotyped BN or EDNOS cases were more likely to be included in this study if they had a lifetime diagnosis of AN or any of the other psychiatric disorders (e.g., schizophrenia, autism) ascertained for the iPSYCH case-cohort study (19,22). In other words, BN and EDNOS cases without other lifetime psychiatric diagnoses (included in the iPSYCH case-cohort study) are not captured in our sample. Therefore, our findings might not be generalizable to all BN or EDNOS cases. Another limitation is that we are unable to estimate the extent to which hospital treatment may have impacted observed ED transitions and remission. For example, treatment for AN could have led to an increased bodyweight and this could have shifted the diagnosis from AN to EDNOS despite the presence of core AN symptoms. The majority of individuals in this study are of European ancestry making the results of this study less generalizable to other non-European populations.

In conclusion, this study supports previous findings that the majority of ED patients do not experience diagnostic transitions and that presumed remission is likely regardless of ED diagnosis. This study highlights potential genetic influences underlying ED transitions and remission but also points towards a possible unique genetic architecture that is not captured by our selection of PGS. Further research on understanding possible environmental influences on ED transitions and remission is warranted as this may give us a deeper understanding of the etiology of EDs and inform tertiary prevention strategies that could lead to improved outcomes.

## Data Availability

Access to data requires application to the Danish Health Data Authority and the Danish Data Protection Agency.

## Acknowledgements

The study was supported by the Novo Nordisk Foundation (grant no. NNF20OC0064993). The iPSYCH data was supported by grants from the Lundbeck Foundation (grant no. R102-A9118, R155-2014-1724, and R248-2017-2003) and the Universities and University Hospitals of Aarhus and Copenhagen. The Anorexia Nervosa Genetics Initiative (ANGI) was an initiative of the Klarman Family Foundation, further anorexia nervosa genotype data were supported by grant from the Lundbeck Foundation (grant no. R276-2018-4581). Mohamed Abdulkadir acknowledges grant support from the National Institute of Mental Health (R01MH120170). Zeynep Yilmaz acknowledges grant support from the Independent Research Fund Denmark (DFF, Sapere Aude, grant no. 1052-00029B). CMB is supported by NIMH (R56MH129437; R01MH120170; R01MH124871; R01MH119084; R01MH118278; R01 MH124871); Swedish Research Council (Vetenskapsrådet, award: 538-2013-8864); Lundbeck Foundation (Grant no. R276-2018-4581). Bjarni J. Vilhjalmsson was supported by a grant from the Lundbeck Foundation (R335-2019-2339), and a grant from the Independent Research Fund (2034-00241B).

