## Supplemental Methods for "Descriptives and genetic correlates of eating disorder diagnostic transitions and presumed remission in the Danish registry"

*Definition of distinct eating disorder (ED) episodes*

From the in- and outpatient contacts in the National Patient Register (NPR) and the Psychiatric Central Research Register (PCRR) for each individual, distinct episodes of an ED were defined (see Figure S1 for a diagram of how the ED episodes were defined). As a first step, we combined records of the patients into contacts, keeping in mind the following conditions. When the records indicated different ED diagnosis types (e.g., main diagnosis, secondary diagnosis) during the same contact period, the main diagnosis was chosen over the secondary diagnosis or the underlying auxiliary diagnosis. In the situation of multiple records during the same contact period, such as a diagnosis from a psychiatric ward and a diagnosis from a somatic ward, the diagnosis from the psychiatric ward was chosen. In the case of multiple records with different main ED diagnoses during the same contact, the AN diagnosis trumped a BN diagnosis, and a BN diagnosis trumped an EDNOS diagnosis. When concurrent inpatient and outpatient records existed for the same patient during the same period, the diagnosis recorded in the inpatient record was used.

We then further organized the contacts into distinct episodes of an ED. Instances of overlapping contacts or immediately adjacent contacts of the same patient type (e.g., inpatient) and same diagnosis were combined into one episode, with the start date of the episode set at the initiation of the earlier contact and the end date set at the end of the later contact. Situations wherein inpatient contacts were contained within a longer outpatient contact were resolved by merging the two contacts into one episode, choosing the outpatient diagnosis. Instances of short contacts in proximity (within 30 days) to a longer contact were merged into one episode, and differences in diagnosis were resolved by choosing the diagnosis of the longer contact. In situations where a short contact was equally close (within 30 days) to a preceding longer contact and a succeeding longer contact, the short contact was merged with the preceding earlier contact into one episode, with diagnostic differences being resolved by choosing the diagnosis of the longer contact. Multiple short contacts in proximity (within 30 days) were merged into one episode, and conflicts in diagnosis was resolved by choosing AN diagnosis over a BN and choosing a BN diagnosis over a diagnosis of EDNOS. We kept occurrences of long contacts in proximity (within 30 days) and short contacts not in proximity as separate episodes.

The organization of the records into contacts and into episodes resulted in three possible ED episodes: AN; BN; and EDNOS. We also included a state of presumed remission; individuals that were contact free regarding their ED for at least 2 years.

**Filtering of PGS**

From the 940 PGSs calculated using LDpred2, we performed pre-filtering on the PGS prior to association analysis based on the following criteria (for a flowchart refer to Figure S2): (i) heritability of the trait could not be determined (N = 2); (ii) the heritability estimate from LDpred2 differed significantly (i.e., more than three standard deviations) from the heritability estimate from linkage disequilibrium score regression(1) (n = 19); (iii) heritability estimate was less than 0.01 (n = 37); (iv) duplicated PGSs (one of each duplicated pair was removed; n = 32); or (v) PGSs were highly correlated (from each PGS pair with r ≥ 0.75, the PGS that on average had higher correlations with the rest of the PGSs in the dataset was removed, resulting in the removal of 223 PGSs). Furthermore, a subset of UK Biobank (UKBB) PGSs underwent additionally filtering (n = 211) to evaluate biological and genetic relevance to our outcomes of interest, especially given that the UKBB demographic encompasses an older population (average age = 56.52 years)(2). Specifically, we decided to remove UKBB derived PGSs in the domains of self-reported diet traits (e.g., type of cereal bran eaten, consumption of instant coffee, milk type used), familial history traits (e.g., paternal history of heart disease, maternal history of lung cancer), aging traits (e.g., pattern of baldness, facial ageing, eye problems), lifestyle traits (e.g., exposure to loud music, reason for discontinuing smoking), medication traits (e.g., use of Ibuprofen, Lisinopril, Ramipril), as these are domains that are very likely to be associated with age (i.e., diet and medication use) and may also suffer from recall bias (e.g., family history). Following filtering, a total of 422 PGSs were included in our analyses (Table S1). All PGSs were standardized to a mean of 0 and SD of 1.
