## Supplemental Figures S1 S2 for "Descriptives and genetic correlates of eating disorder diagnostic transitions and presumed remission in the Danish registry"

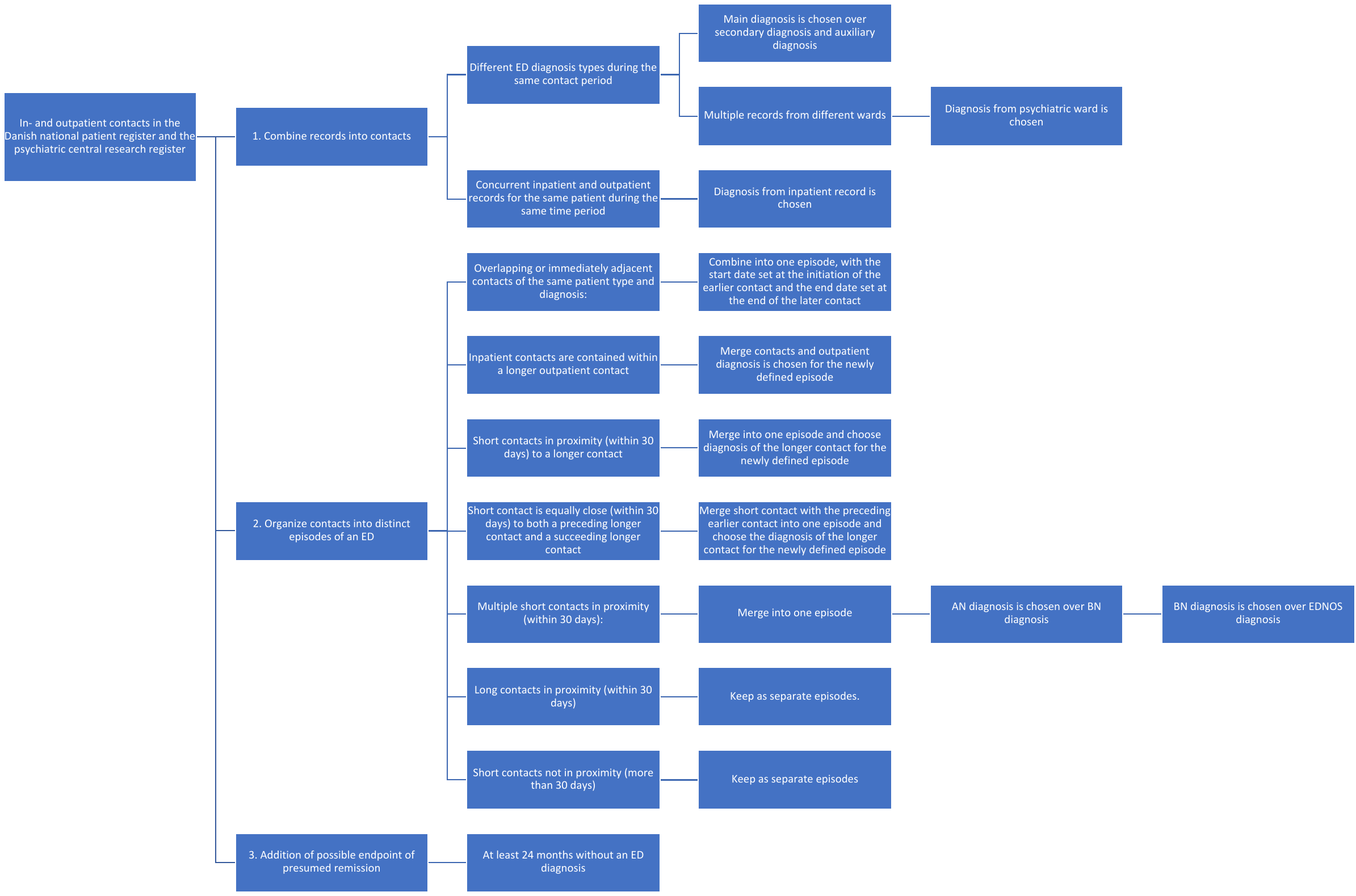

**Figure S1:** Diagram depicting how distinct eating disorder (ED) episodes were defined using records from the Danish National Patient Register and the Psychiatric Central Research Register.

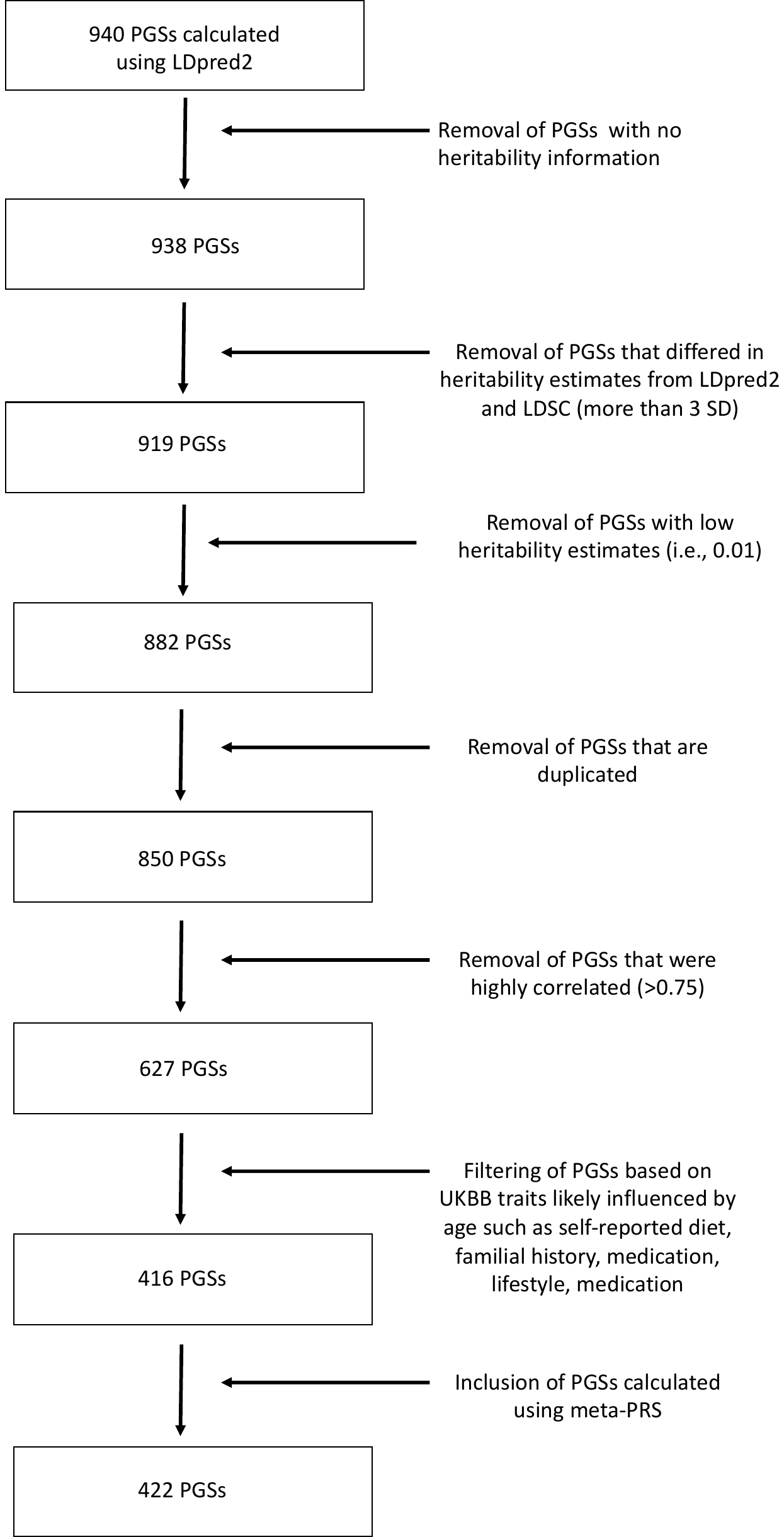

**Figure S2:** Flowchart depicting filtering of polygenic scores (PGS) calculated with LDpred2 (1) and meta-PRS (2) included in the final analysis. SD, standard deviation. UKBB, UK Biobank (3)
